# Rapid Acquisition and Transmission of Drug Resistance Amongst Beijing Lineage *Mycobacterium tuberculosis* in Vietnam

**DOI:** 10.1101/2022.11.03.22281872

**Authors:** Matthew Silcocks, Xuling Chang, Nguyen Thuy Thuong Thuong, Youwen Qin, Dang Thi Minh Ha, Phan Vuong Khac Thai, Srinivasan Vijay, Do Dang Anh Thu, Hoang Ngoc Nhung, Nguyen Huu Lan, Nguyen Thi Quynh Nhu, David Edwards, Artika Nath, Kym Pham, Nguyen Duc Bang, Tran Thi Hong Chau, Guy Thwaites, A. Dorothee Heemskerk, Chiea Chuen Khor, Yik Ying Teo, Michael Inouye, Rick Twee-Hee Ong, Maxine Caws, Kathryn E. Holt, Sarah J. Dunstan

## Abstract

Whole genome sequencing (WGS) and phenotypic drug susceptibility testing was performed on a collection of 2,542 *Mycobacterium tuberculosis (Mtb)* isolates from tuberculosis (TB) patients recruited in Ho Chi Minh City (HCMC), Vietnam, to investigate *Mtb* diversity, the prevalence and phylodynamics of drug resistance, and *in silico* resistance prediction with sequencing data. Amongst isolates tested phenotypically against first-line drugs, we observed high rates of streptomycin [STR, 37.7% (N=573/1,520)] and isoniazid resistance [INH, 25.7% (N=459/1,786)], and lower rates of resistance to rifampicin [RIF, 4.9% (N=87/1,786)] and ethambutol [EMB, 4.2% (N=75/1,785)]. Resistance to STR and INH was predicted moderately well when applying the TB-Profiler algorithm to WGS data (sensitivities of 0.81 and 0.87 respectively), while resistance to RIF and EMB was predicted relatively poorly (sensitivities of 0.70 and 0.44 respectively). Rates of multidrug-resistance [(MDR, 3.9% (N=69/1,786)], and resistance to a number of second-line drugs [Para-aminosalicylic acid (29.6% N=79/267), Amikacin (15.4% N=41/267) and Moxifloxacin (21.3%), N=57/267], were found to be high within a global context. Comparing rates of drug resistance among lineages, and exploring the dynamics of resistance acquisition through time, suggest the Beijing lineage (lineage 2.2) acquires *de novo* resistance mutations at higher rates and suffers no apparent fitness cost acting to impede the transmission of resistance. We infer resistance to INH and STR to have arisen earlier, on average, than resistance to RIF, and to be more widespread across the phylogeny. The high prevalence of ‘background’ INH resistance, combined with high rates of RIF mono-resistance (20.7%, N=18/87) suggests that rapid assays for INH resistance will be valuable in this setting. These tests will allow the detection of INH mono-resistance, and will allow MDR isolates to be distinguished from isolates with RIF mono-resistance.

## Introduction

Tuberculosis (TB) remains a global epidemic with one quarter of the world’s population estimated to be infected. The impact of the COVID-19 pandemic on essential TB services has reversed years of progress, with the number of newly diagnosed TB patients falling to 5.8 million in 2020, much less than the estimated 10 million who developed TB [Global Tuberculosis Report, 2021 (WHO, 2021)]. An increase in TB deaths in 2020 was also estimated (1.3 million in HIV negative, and 214,000 in HIV-positive) due largely to a reduction in the number of people treated for drug resistant TB [Global Tuberculosis Report, 2021 (WHO, 2021)]. Geographically, the burden of disease lies mainly in South-East Asia (44% of TB cases in 2018) [Global Tuberculosis Report, 2019 (WHO, 2019)], with 86% of new TB cases worldwide being reported from 30 high TB burden countries in 2020 [Global Tuberculosis Report, 2021 (WHO, 2021)].

Vietnam is a high TB burden country, designated by its high number of incident TB and multi-drug resistant TB (MDR-TB) cases [WHO global lists of high burden countries for TB, multidrug/rifampicin-resistant TB (MDR/RR-TB) and TB/HIV, 2021–2025, (WHO, 2021)]. In 2018, 174,000 and 8,600 people in Vietnam fell ill with TB and drug-resistant TB, respectively. Of the patients with drug-resistant TB, only 36.3% were laboratory confirmed and 36.2% started on second-line treatment.

Although the COVID-19 pandemic has set back recent progress, steps towards the WHO END-TB targets have been made. To be able to achieve a 95% reduction of TB deaths by 2035 (The END TB strategy, World Health Organisation, 2015), modern technologies must be embraced to find innovative ways to accelerate TB control and elimination. One such technology is genomic sequencing, which offers a myriad of opportunities for innovation in diagnostics, treatment, prevention and control of TB.

Genotype data, for example, can be used to predict drug resistance in *Mtb* isolates, by querying databases of confirmed or suspected resistance conferring variants (CRyPTIC Consortium et al. 2018; Walker et al. 2015; Walker et al. 2022). This approach provides an efficient alternative to traditional phenotypic methods which are prone to human error, contamination, and require time-consuming bacterial culturing (Zignol et al. 2018; Gygli et al. 2019; Walker et al. 2022). This technology has the capacity to reduce the probability of misassignment of drugs to patients infected with resistant *Mtb* (compared to standardised drug regimens), and to reduce the time before patients receive effective treatment, potentially leading to more favourable treatment outcomes (Iketleng et al. 2018).

Despite the promise of genotype-based drug resistance prediction, its accuracy has been shown to vary according to human population, *Mtb* lineage, type of drug, and the prediction protocol used (Papaventsis et al. 2017). A more comprehensive understanding of the effect these factors have on prediction accuracy, and the levels of accuracy which are attainable across diverse cohorts, is necessary prior to implementation of these test in all clinical settings.

Genomics also provides insight into the emergence, transmission and overall dynamics of drug resistance in *Mtb*, via the use of a phylogenetic toolkit (Eldholm et al. 2015; Woolenberg et al. 2017; Brown et al. 2019; Cohen et al. 2019; Casali et al. 2014; Manson et al. 2017). Prior studies have produced time-calibrated phylogenies to date the acquisition of drug resistance in *Mtb* lineages (Eldholm et al. 2015; Cohen et al. 2015), and explored their expansion through time (Brown et al. 2019). Others have used models of ancestral sequence reconstruction (Woolenberg et al. 2017; Cohen et al. 2015; Manson et al. 2017) and SNP clustering methods (Casali et al. 2014; Yang et al. 2017) to compare the rates of acquired (*de novo*), versus primary (i.e. transmitted) drug resistance within a population.

While these studies naturally vary in their scope, cohort size, geographic scale and setting, their results have highlighted similar trends in drug resistance evolution. They show that the global drug resistance burden has arisen through both the *de novo* acquisition of drug resistance, and through the transmission of drug resistant *Mtb* to new hosts (Woolenberg et al. 2017; Casali et al. 2014; Ektefaie et al. 2021; Yang et al. 2017; Mai et al. 2018). They also show consistent trends in the order of drug resistance acquisition, with INH resistance generally arising prior to RIF resistance, and being more deeply rooted in the phylogeny (Cohen et al. 2015; Ektefaie et al. 2021; Manson et al. 2017). Finally, when characterising *Mtb* lineage diversity, these studies have implicated Beijing lineage isolates in many outbreaks (Woolenberg et al. 2017; Yang et al. 2017; Casali et al. 2014).

Here we apply these tools to investigate resistance evolution in a South East Asian context, using a cohort from HCMC, Vietnam, which has a high frequency of TB, drug resistance, and the Beijing lineage. This study builds on our previous investigation of *Mtb* diversity in HCMC (Holt et al. 2018), by using an expanded isolate collection and resistance phenotype data to explore the prevalence and transmission dynamics of drug resistance, and our ability to predict it *in silico*.

## Results

### Clinical characteristics

To characterise the diversity and drug resistance of *Mtb* in HCMC, Vietnam, we analysed the genomic sequences of N=2,542 isolates (post quality-filtering) cultured from TB patients between 2001-2013. A subset of these genomes (N=1,627), derived from pulmonary TB (PTB) patients, were published previously (Holt et al. 2018). Here these are supplemented with an additional N=914 novel genomes, comprising N=747 from PTB cases and 167 from TBM cases (Heemskerk et al. 2016; Thwaites et al. 2004). A single lineage 5 genome was included as an outgroup for phylogenetic analyses.

PTB patients were sputum culture positive and ≥ 18 years old (Thai et al. 2018) whereas TBM patients were cerebral spinal fluid culture positive and ≥15 years old (Heemskerk et al. 2016; Thwaites et al. 2004). All TB patients were human immunodeficiency virus (HIV)negative. Overall the patients were predominantly male (71.5% male, 28.5% female) with a median (interquartile) age of 39 (26-49) years. We found the frequencies of males and females to differ across clinical phenotypes for PTB cases, with 73.1% of cases being male, and 26.9% being female. We calculated TBM cases to be more evenly distributed across genders (47.9% male and 52.1% female), however noted males to be more frequent in the larger TBM cohort from which these samples were derived (Heemskerk et al. 2016).

### Genetic diversity and *Mtb* lineage

In our HCMC PTB/TBM genome dataset (N=2,542), we found East Asian lineage 2 to be the most prevalent (N=1,615, 63.6%), followed by the East African-Indian Ocean lineage 1 (N=649, 25.5%), the European-American lineage 4 (N=275, 10.8%) and the Delhi Central Asian lineage 3 (N=2, 0.1%; Figure 1). The pattern of *Mtb* genomic diversity in the HCMC PTB/TBM genome dataset was thus consistent with our published subset of Vietnamese genomes (Holt et al. 2018).

**Figure 1.**
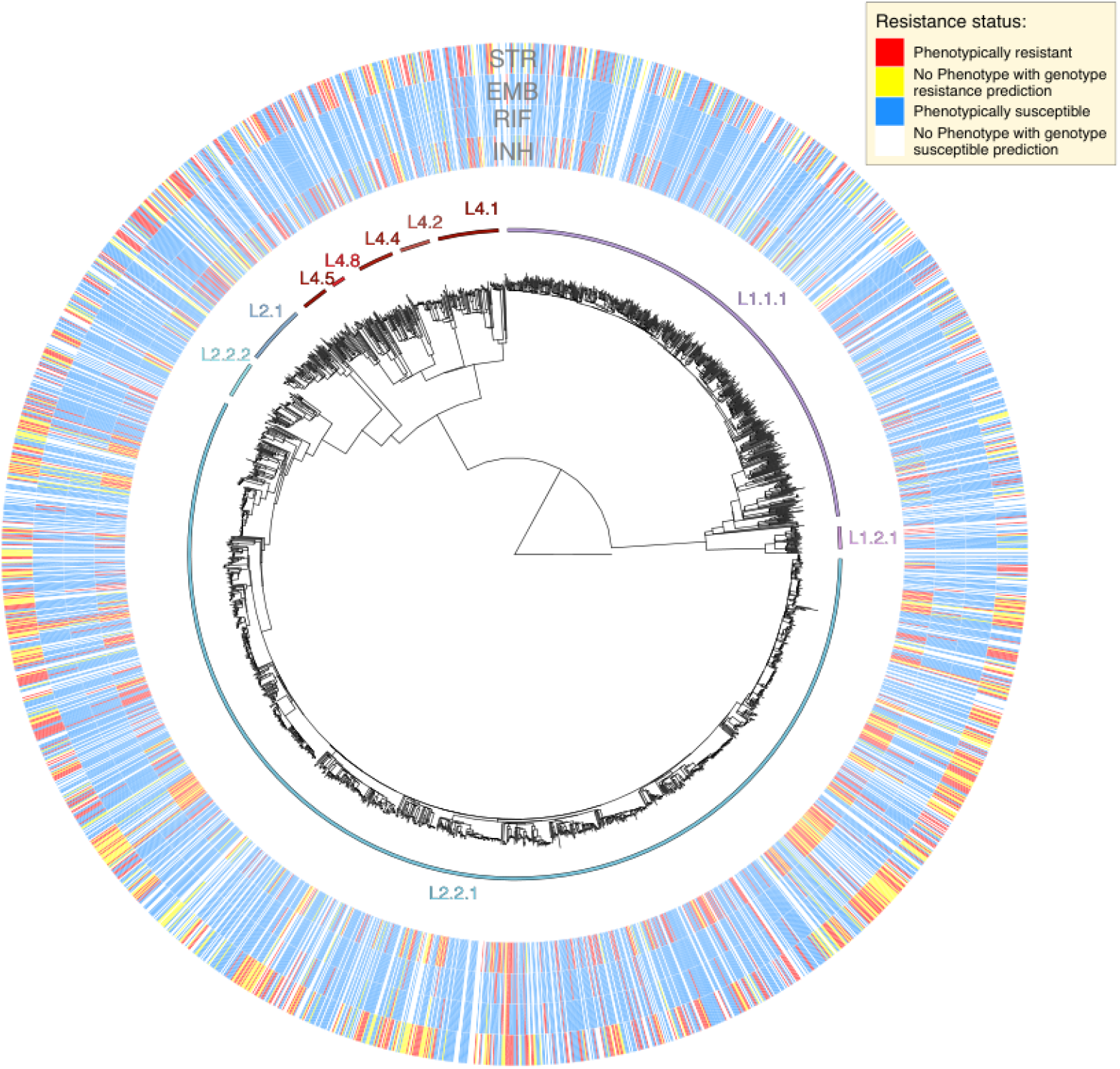
Phylogeny of 2,542 *Mtb* genomes, with associated phenotypic and genotypic drug resistance data for four first line drugs (moving outwards: INH, RIF, EMB and STR, respectively). For all samples with gold-standard phenotypic drug susceptibility testing (DST) data (N= 1,781), red denotes resistant isolates and blue denotes susceptible isolates. For all isolates without phenotypic DST data, yellow denotes genotypically predicted resistant isolates and white denotes genotypically predicted susceptible isolates. As noted elsewhere, phenotypic resistance and genotypic-based resistance prediction statuses were not perfectly correlated. Of all isolates predicted to be resistant using genotype data, 92.6, 79.2, 40.2 and 86.5% were phenotypically resistant for INH, RIF, EMB, STR respectively (positive predictive value; Figure 2). Of all isolates predicted to be susceptible using genotype data, 95.6, 98.5, 97.6 and 89.2% were phenotypically susceptible for these same four drugs (negative predictive value; Figure 2).

To gain a more nuanced understanding of patterns of *Mtb* diversity in our genome dataset, we also considered lineage distributions at a more granular level of phylogenetic resolution. While we recognise that several recent studies have proposed new *Mtb* lineage nomenclature schemes (Napier et al. 2020; Freschi et al. 2021; Gisch et al. 2022; Palittapongarnpim et al. 2018), here we decided to define lineages according to the most detailed level of resolution described by Coll et al. (2014), until a consensus is established, and a clearer understanding of sublineage distributions under these new schemes is obtained.

Consistent with our prior survey (Holt et al. 2018), lineage 1 isolates mainly belonged to sublineages 1.1.1.1 (N=525) and 1.1.1 (N=83), which predominate in Mainland South East Asia (Netikul et al. 2021; Freschi et al. 2021; Figure 1). We also observed isolates belonging to sublineage 1.2.1 (N=33), which attains highest frequencies in Island South East Asia (Netikul et al. 2021; Menardo et al. 2021).

Isolates from lineage 2 could be partitioned into both the ‘Proto-Beijing’ (L2.1; N=86) and Beijing (L2.2; N=1,529) sublineages (Luo et al. 2015; Figure 1). Beijing lineage isolates belonged to both L2.2.1 (N=1,446), which has a wide global distribution (Holt et al. 2018), as well as the less prevalent L2.2.2 (N=55) (Shiticov et al. 2017). We documented a wide array of L4 sublineages, consistent with a history of repeated introductions into Southeast Asia, as inferred by Holt *et al*. (2018). The most frequent L4 sublineages were 4.1, 4.2, 4.4 and 4.5.

Also consistent with Holt *et al*. (2018), we found *Mtb* lineage to be significantly associated with both the sex (Table 1), and age of the patient. While patients were predominantly male, lineage 2 and sublineage 2.2.1 were more prevalent amongst female patients (lineage 2;

**Table 1.** Lineage distribution by sex and clinical phenotype.

|  |  | Lineage |  |  |  |  |  |  |
| --- | --- | --- | --- | --- | --- | --- | --- | --- |
|  |  | 1 | 2.1 | 2.2.1 | 2.2 (Other than 2.2.1) | 3 | 4 | total |
| Sex | Male | 496 (27.3%) | 68 (3.7%) | 1,014 (55.8%) | 44 (2.4%) | 1 (0.1%) | 193 (10.6%) | 1,816 (71.5%) |
|  | Female | 153 (21.1%) | 18 (2.5%) | 452 (62.3%) | 19 (2.6%) | 1 (0.2%) | 82 (11.3%) | 725 (28.5%) |
| Clinical Phenotype |  |  |  |  |  |  |  |  |
|  | PTB | 581 (24.5%) | 82 (3.50%) | 1,389 (58.5%) | 57 (2.4%) | 2 (0.1%) | 263 (11.1%) | 2,374 (93.4%) |
|  | TBM | 68 (40.7%) | 4 (2.4%) | 77 (46.1%) | 6 (3.6%) | - | 12 (7.2%) | 167 (6.6%) |
|  | all TB | 649 (25.5%) | 86 (3.4%) | 1,466 (57.7%) | 63 (2.5%) | 2 (0.1%) | 275 (10.8%) | 2,541 |

67.4% of females vs 62.0% of males; *p*=0.010), (sublineage 2.2.1; 62.3% of females vs 55.8% of males; *p*=0.005 (when excluding other L2 isolates from test)). Lineage 1 strains showed higher prevalence amongst males (27.3% of males vs 21.1% of females; *p*=0.001; Table 1). Lineage 2 was significantly associated with younger people (median 37 years), and lineage 1 with older people (median 43 years, *p*=8.36×10^−8^).

### Phenotypic drug resistance of PTB isolates

Phenotypic drug susceptibility testing was performed for a subset of the sequenced isolates (N=1,786). We assessed phenotypic resistance to three first line drugs (INH, RIF and EMB) using MGIT (N=1,786), and also tested a subset of these isolates using UKMYC5 (N=267). Isolates were deemed resistant to an antimicrobial if they were classified as resistant by one or both methods. In total, N=459/1,786 (25.7%) isolates were resistant to INH, N=87/1,786 (4.9%) to RIF, and N=75/1785 (4.2%) to EMB. In total, 3.9% (N=69/1,786) were MDR (INH and RIF resistant; Table 2), and 20.7% (N=18/87) of RIF resistant isolates were INH sensitive. STR resistance, which was tested by MGIT only, was identified in 37.7% (573/1,520) of the tested isolates. The frequencies of resistance to second-line antibiotics, determined for 267 isolates only, are given in Table 2.

**Table 2.** Antimicrobial resistance by lineage.

| Drug resistance in PTB | 1 | 2.1 | 2.2.1 | 2.2 (Other than 2.2.1) | 4 | total resistant | total tested |
| --- | --- | --- | --- | --- | --- | --- | --- |
| **MDR** |  |  |  |  |  |  |  |
| Isoniazid/Rifampicin | 7 (10.1%) | 4 (5.8%) | 53 (76.8%) | 2 (2.9%) | 3 (4.3%) | 69 (3.9%) | 1,786 |
| **Antimicrobial (DST by either)** |  |  |  |  |  |  |  |
| Isoniazid | 74 (16.1%) | 18 (3.9%) | 311 (67.8%) | 6 (1.3%) | 50 (10.9%) | 459 (25.7%) | 1,786 |
| Rifampicin | 11 (12.6%) | 5 (5.7%) | 64 (73.6%) | 2 (2.3%) | 5 (5.7%) | 87 (4.9%) | 1,786 |
| Ethambutol | 7 (9.3%) | 2 (2.7%) | 58 (77.3%) | 1 (1.3%) | 7 (9.3%) | 75 (4.2%) | 1,785 |
| **Antimicrobial (DST by MGIT)** |  |  |  |  |  |  |  |
| Streptomycin | 78 (13.6%) | 29 (5.1%) | 391 (68.2%) | 11 (1.92%) | 64 (11.2%) | 573 (37.7%) | 1,520 |
| **Antimicrobial (DST by UKMYC5)** |  |  |  |  |  |  |  |
| Bedaquiline (BDQ >=1) | - | - | - | 1 (100%) | - | 1 (0.4%) | 267 |
| Kanamycin (KAN>=2.5) | 1 (6.7%) | 2 (13.3%) | 5 (33.3%) | 1 (6.7%) | 6 (40.0%) | 15 (5.6%) | 267 |
| Ethionamide (ETH>=5) | 12 (54.5%) | - | 10 (45.5%) | - | - | 22 (8.2%) | 267 |
| Amikacin (AMI>=1) | 10 (24.4%) | 1 (2.4%) | 22 (53.7%) | 4 (9.8%) | 4 (9.8%) | 41 (15.4%) | 267 |
| Levofloxacin (LEV >=1) | 8 (25.8%) | 1 (3.2%) | 18 (58.1%) | 2 (6.5%) | 2 (6.5%) | 31 (11.6%) | 267 |
| Moxifloxacin (MXF >=0.5) | 10 (17.5%) | 1 (1.8%) | 33 (57.9%) | 5 (8.8%) | 8 (14.0%) | 57 (21.3%) | 267 |
| Delamanid (DLM >= 0.12) | - | - | 4 (80.0%) | 1 (20.0%) | - | 5 (1.9%) | 267 |
| Linezolid (LZD >=1) | 14 (43.8%) | - | 13 (40.6%) | - | 5 (15.6%) | 32 (12.0%) | 267 |
| Ceftazadime (CFZ >=1) | 2 (28.6%) | - | 3 (42.9%) | - | 2 (28.6%) | 7 (2.6%) | 267 |
| Rifabutin (RFB >=1) | - | 1 (20.0%) | 4 (80.0%) | - | - | 5 (1.9%) | 267 |
| Para-aminosalicylic acid (PAS>=4) | 31 (39.2%) | 2 (2.5%) | 36 (45.6%) | 5 (6.3%) | 5 (6.3%) | 79 (29.6%) | 267 |

We noted that rates of resistance to all first line drugs were higher amongst Beijing lineage isolates (L2.2) than isolates belonging to lineages 1 and 4 [Pearson’s Chi-Squared test; INH (*p*=2.07×10^−4^), RIF (*p*=0.003), EMB (*p*=0.002) and STR (*p*=8.29×10^−12^)]. This finding was in keeping with several prior studies (Casali et al. 2014; Borrell et al. 2009; Neimann et al. 2010; Woolenberg et al. 2017).

### Prediction of drug resistance using genotype data

We assessed our ability to correctly predict drug resistance using genotype data, and found it to vary according to the drug, and the phenotype (either ‘resistant’ or ‘susceptible’) being predicted. We analyzed raw sequencing data with TB-Profiler (Phelan et al. 2019), and calculated sensitivity and specificity metrics using standard approaches (Mahe et al. 2019). Sensitivity, which measure the proportion of phenotypically resistant strains correctly predicted to be resistant, for first line drugs was 0.87, 0.70, 0.44 and 0.81 for INH, RIF, EMB and STR respectively (Figure 2a). Specificity, which measure the proportion of phenotypically susceptible strains correctly predicted to be susceptible, was 0.98, 0.99, 0.97 and 0.92 for these same four drugs respectively (Figure 2a).

**Figure 2.**
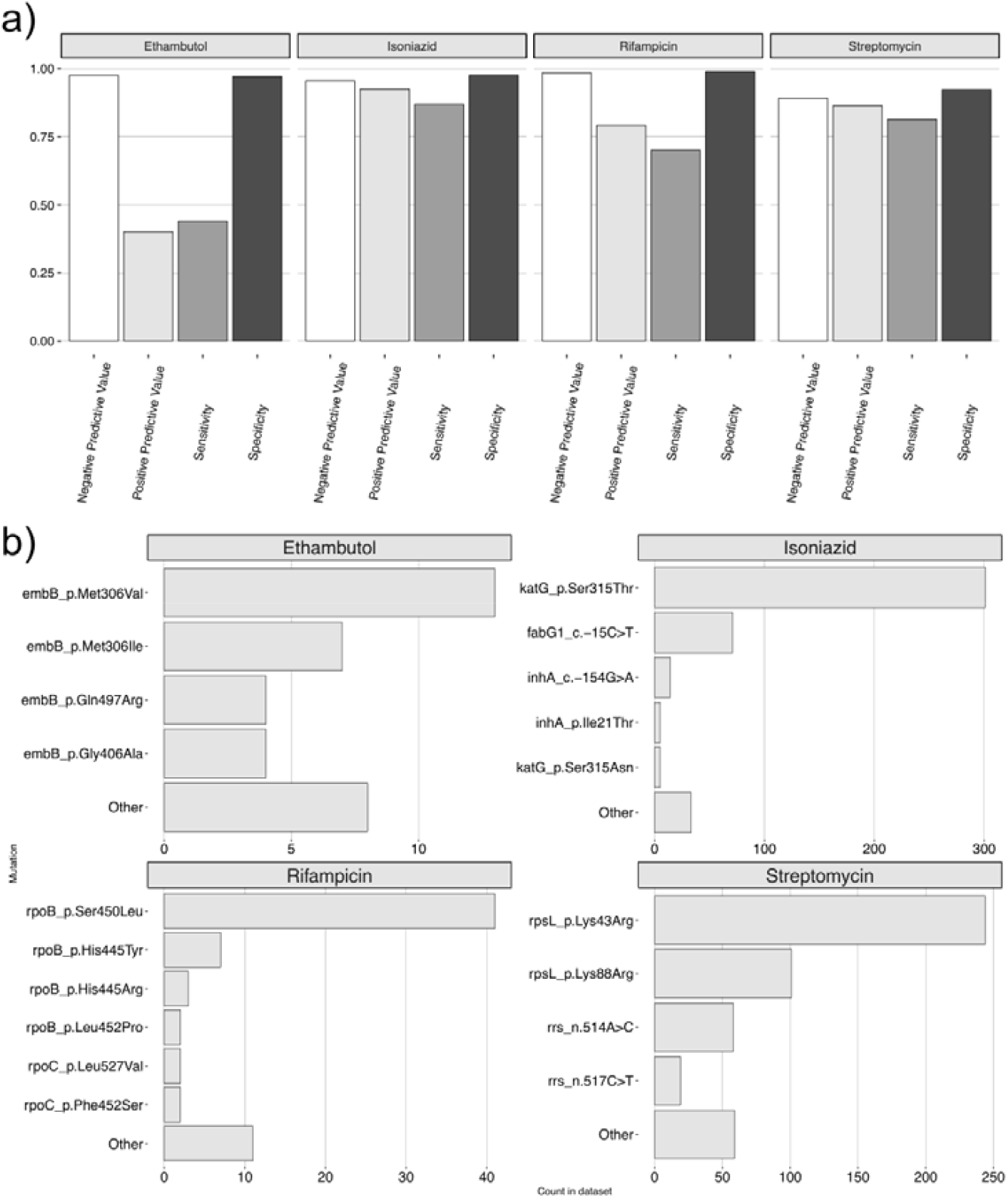
**a)** Sensitivity, specificity, positive and negative predictive values for four first line drugs inferred using TB-Profiler. **b)** The count of resistance conferring mutations/amino acid substitutions in phenotypically resistant isolates.

Positive predictive values, which measure the proportion of isolates with a resistance prediction which were phenotypically resistant, were 0.93, 0.79, 0.40 and 0.87 for INH, RIF, EMB and STR (Figure 2a). Negative predictive values, which measure the proportion of isolates with a susceptible prediction which were phenotypically susceptible were 0.96, 0.98, 0.98 and 0.89 (Figure 2a).

The frequency distribution of mutations explaining resistance to first-line drugs (Figure 2b) was skewed towards a number of commonly reported variants (WHO, 2021a). In particular, the katG-Ser315Thr substitution was most frequently observed in INH resistant cases (N=299/459, 65.1% of resistant isolates), followed by fabG1-15C>T (N=71/459, 15.5%). RIF resistance was most commonly explained by rpoB-Ser450Leu (N=41/87, 47.1%), and STR resistance by rpsL-Lys43Arg (N=243/573, 42.4%) and rpsL-Lys88Arg (N=100/573, 17.5%). EMB displayed a wider spectrum of resistance conferring mutations, with the most frequent being embB-Met306Val (N=13/75, 17.3%) and embB-Met306Ile (N=7/75, 9.3%). Importantly, in the majority of cases (N=42/75, 56%), resistance to EMB was not explained by any known markers.

Resistance prediction for second-line drugs was overall poor [sensitivity values: BDQ=0 (N=1 resistant isolate), ETH=0.41 (N=22), AMI=0 (N=41), LEV=0.07 (N=31), MXF=0.04 (N=57), DLM=0.8 (N=5), LZD=0 (N=32), CFZ=0 (N=7), BDQ=0 (N=1), KAN=0 (N=15) and PAS=0.05 (N=79)].

We found the frequency of putative resistance-conferring variants to be similar between TBM and PTB isolates (30.5% vs 24.9% for INH, 1.2% vs 4.7% for RIF, 3.0% vs 4.9% for EMB, 40.1% vs 36.4% for STR). We further verified that TBM isolates were dispersed throughout the phylogeny, and were not clustered in monophyletic clades (supp fig S1), justifying their inclusion in the subsequent analysis of antibacterial resistance dynamics.

### Dynamics of drug resistance acquisition and transmission

We investigated the dynamics of drug resistance evolution within our cohort, to understand temporal and lineage-specific trends in drug resistance development, and to characterise the relative burden of acquired versus transmitted resistance. To do this, we used ancestral state reconstruction to map individual resistance mutation events to the phylogeny and identify instances of transmitted (Figure 3a; red points. see Methods) and acquired resistance (Figure 3a; blue points).

**Figure 3.**
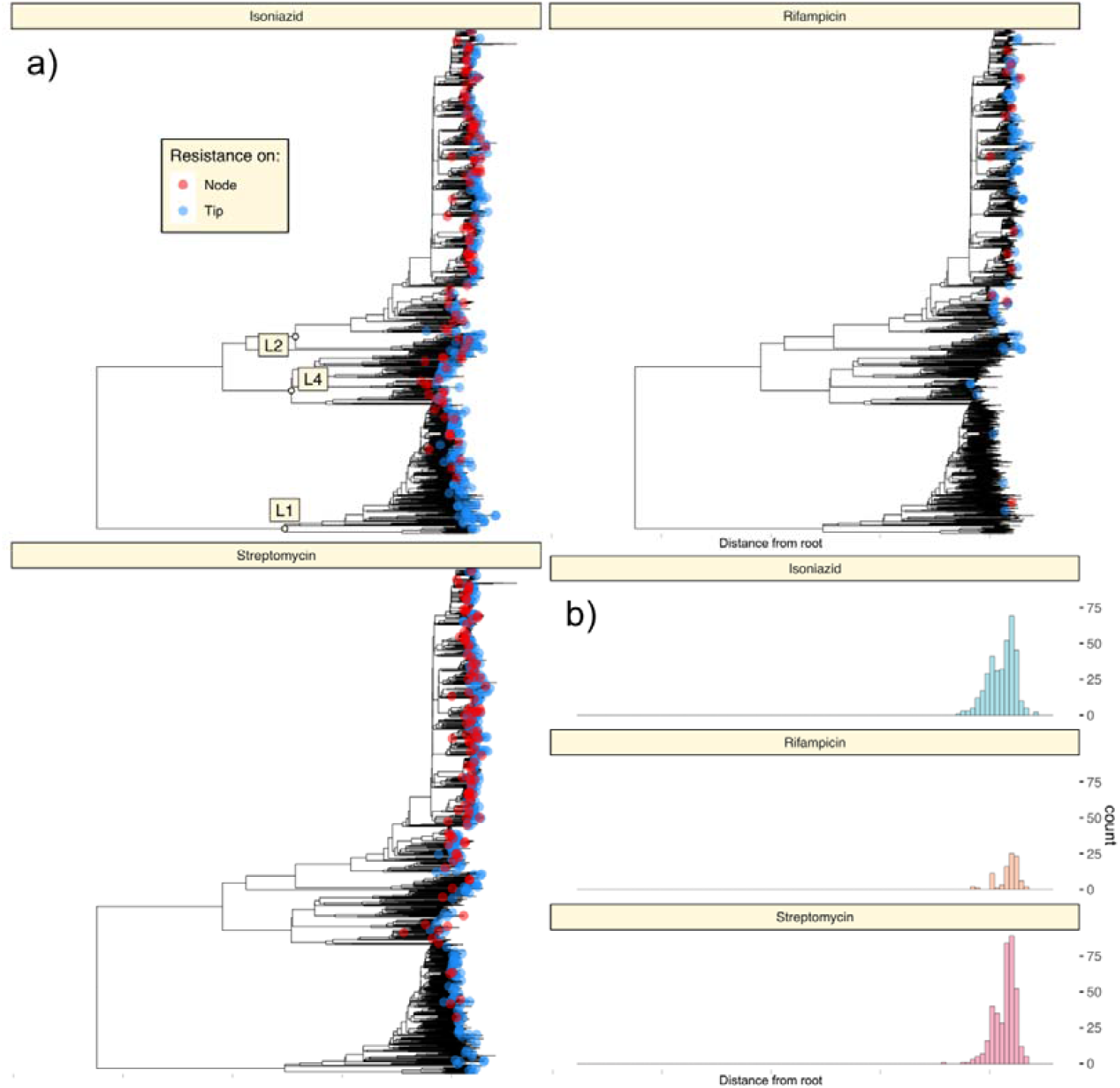
**a)**. Acquisition and transmission of mutations conferring resistance to three first line drugs: INH, RIF and STR. Biallelic SNPs present within the TB-Profiler catalogue were mapped onto nodes of the phylogeny using SNPPar and labelled according to whether they arose on a tip/terminal branch (blue circles), or interior branch (red circles). b) Histograms showing the distribution of resistance mutation emergence events relative to the root of the tree. X-axis scale of the histogram matches that of the above phylogeny.

Across the phylogeny, we inferred 826 unique mutation events leading to INH, STR or RIF resistance (EMB was not modelled due to the low positive predictive power of known variants, see Fig 2a). We observed a trend in the order of resistance acquisition (Figure 3), with INH and STR mutations arising earlier (closer to the root of the tree) than RIF mutations (Figures 3a and b; median heights of 0.01285, 0.01291 and 0.01299 for INH, STR and RIF variants, respectively). This inference of early and widespread INH resistance was consistent with the conclusions of prior studies (Ektefaie et al. 2021; Manson et al. 2017).

We estimated the relative burden of transmitted (as opposed to acquired) resistance by observing the relative density of resistance mutations mapped to nodes, as opposed to terminal branches of the phylogeny (see Methods). Across the entire tree, we find that 63.6% (402/632) of isolates with INH resistance were categorised as cases of transmitted resistance (i.e. with their resistance mutation mapping to a node of the phylogeny). A similar percentage was calculated for STR variants (66.6%; 519/779).

These rates, while clearly influenced by the design and density of the sampling regime, indicate that DR-TB burden in HCMC is explained by both *de novo* acquisition, and transmission of resistance. As this, and our previous analysis of *Mtb* genetic variation within Vietnam (Holt et al. 2018) sample only a limited fraction of incident TB cases in HCMC, these estimates of transmission rates are likely to be conservative.

### Lineage specific trends in drug resistance evolution

We observed clear lineage-specific trends in the dynamics of drug resistance evolution. Consistent with prior estimates of drug resistance rates across lineages (Casali et al. 2014; Borrell et al. 2009; Neimann et al. 2010; Woolenberg et al. 2017), we observed lineage 2 isolates to acquire resistance conferring mutations at a faster rate than isolates from other lineages. 1.13% of all mutation events across the lineage 2 clade were resistance conferring: a higher figure than for lineages 1 and 4 (0.35% and 0.32% respectively).

Lineage 2 isolates were also found to contribute disproportionately to the number of transmitted resistance cases within our cohort. Of the total 402 cases of transmitted INH resistance across the phylogeny, 347 (86.3%) of these occurred within the lineage 2 subtree, and 20 (5.0%) and 35 (8.7%) occurred within the lineage 1 and 4 subtrees respectively. These proportions were higher than expected when considering the overall lineage distribution across all isolates in the phylogeny (25.5% L1, 63.5% L2, 10.8% L4). Similar findings were reported when considering STR resistance variants (487 of 519 (93.8%) cases of transmitted resistance occurred within the lineage 2 subtree). Thus, lineage 2 isolates possessed both a higher absolute rate of resistance acquisition, and a higher propensity to transmit resistance.

We also found a greater proportion of resistant lineage 2 isolates could be attributed to transmission when compared to other lineages. 74.6% (N=347/465) of INH resistant lineage 2 isolates were classified as cases of transmitted resistance, a higher figure than for lineages 1 (20%; N=20/100) and 4 (52.2%; N=35/67). Similar results were observed when considering STM resistance (L2 – 71.9%; N=487/677, L1 – 15.9%; N=11/69, L4 – 63.6%; N=21/33).

Finally, we investigated whether *Mtb* isolates with genotypic resistance were less likely to be transmitted than susceptible isolates. We are interested in this question, as resistance conferring variants are typically associated with fitness costs in *Mtb*, and other bacterial species (Alame Emane et al. 2021; Nguyen et al. 2018; Gagneux et al. 2009; Andersson & Hughes, 2010). Determining the potential of resistant isolates to transmit to new human hosts will therefore be relevant in determining future drug resistance trajectories worldwide.

Firstly, similar to prior investigations (Casali et al. 2014; Yang et al. 2017; Mai et al. 2018), we measured the proportion of resistant isolates which were part of a transmission cluster, and compared this to the proportion of susceptible isolates in a transmission cluster (Figure 4a). For this and the below analysis we restricted our focus to sublineage 2.2.1, which has previously been shown to display, on average, shorter terminal branches and higher rates of resistance than other lineages in this setting (Holt et al. 2018). We also restricted our analysis to focus on the two drugs with substantial numbers of resistance mutations across the phylogeny, INH and STR. We found isolates with INH and STR resistance mutations to be significantly less likely to be found in a transmission cluster when using a 5-SNP threshold (Figure 4a; Pearson’s Chi-Squared Test *p*=6×10^−4^ for INH and *p*=0.003 for STR), but found no significant differences when using 10- and 15SNP thresholds (*p*=0.51 and *p*=0.18 for INH; *p*=0.24 and *p*=0.28 for STR).

**Figure 4.**
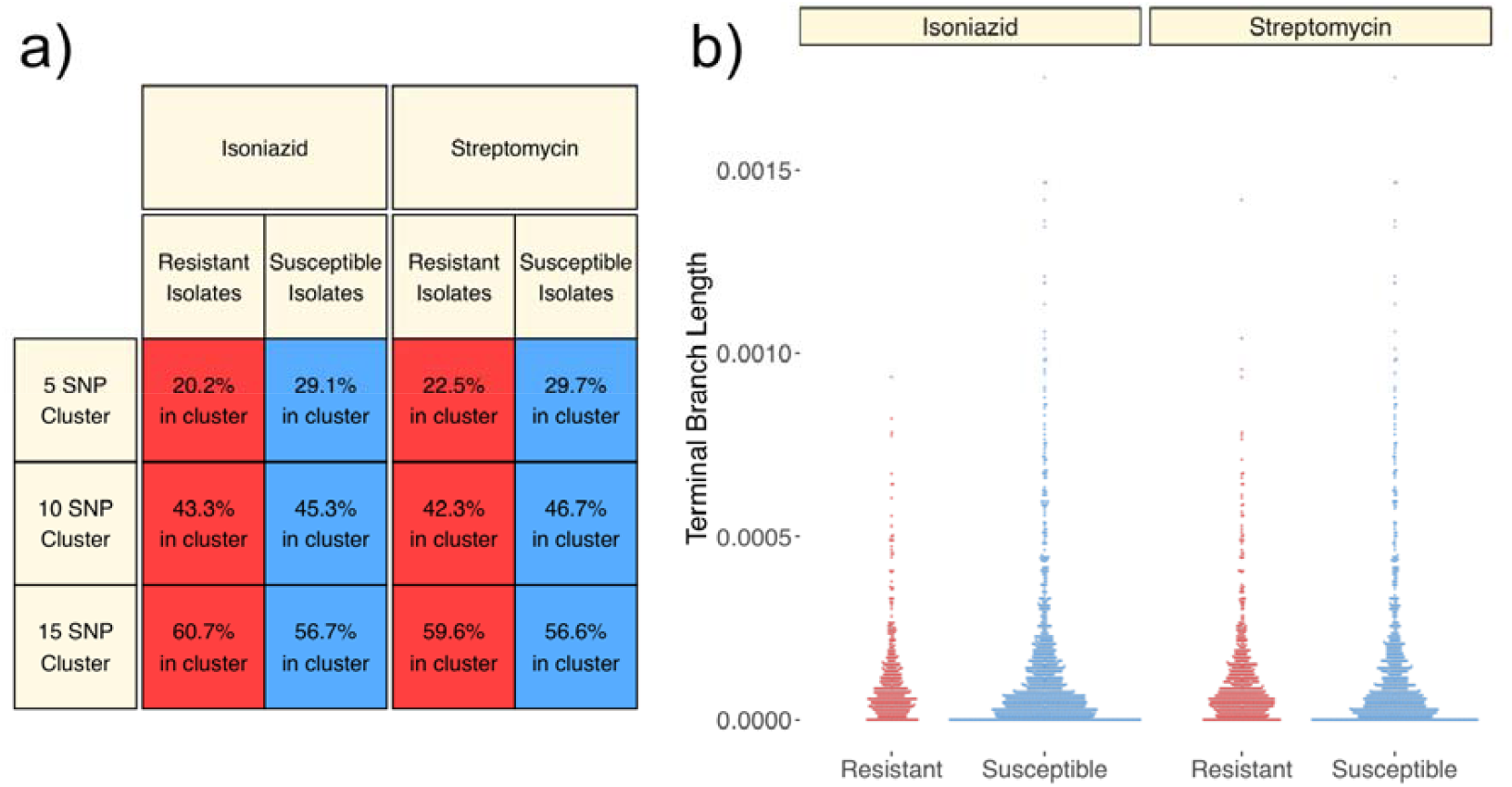
**a)** Percentages of resistant and susceptible L2.2.1 isolates that belong to an epidemiological cluster at 5, 10 and 15 SNP thresholds. Percentages were calculated for INH and STR, the two drugs with the highest rates of resistance. **b)** Beeswarm plots showing the distribution of terminal branch lengths of L2.2.1 isolates leading to tips with INH and STR resistance mutations.

Secondly, as per approaches used in other studies of *Mtb* phylogenetics [(Holt et al. 2018; Freschi et al. 2021) (Figure 4b)], we measured the lengths of terminal branches in the tree leading to resistant and susceptible tips (Figure 4b). We did not find significant differences in the lengths of terminal branches leading to isolates with INH and STR resistance mutations (Figure 4b; Mann Whitney U tests; *p*=0.64 for INH; *p*=0.36 for STR). This evidence suggests that the presence of known drug resistance mutations does not substantially impede the transmission of *Mtb* lineage 2.2.1 isolates from HCMC.

## Discussion

### High prevalence of phenotypic drug resistance in *Mtb* isolates from HCMC

Our investigation has highlighted the high rates of resistance to several first and second line drugs, in addition to MDR, within a Vietnamese cohort. The results reported here are in keeping with a recent national survey of Vietnam (Nhung et al. 2015), which also reports high rates of STR (27.4%) and INH (18.9%) resistance, and lower rates of RIF (4.1%) and EMB (3.4%) resistance amongst new TB cases. This study adds an additional dimension to this survey, by recording rates of phenotypic resistance to second line drugs, and finding rates for PAS, MXF, AMI, LZD and LEV to all be greater than 10%.

When considered globally, individual first line drug resistance values were higher than has been reported in parts of Africa (Ismail et al. 2018; Gehre et al. 2016; Chonde et al. 2010), East Asia (He et al. 2008; Bai et al. 2007), and South Asia (Tahseen et al. 2016), but lower than a South American cohort (Quispe et al. 2020). The second line drug resistance rates we observe are higher than reported in the small number of studies which survey second line resistance in new TB cases (Quispe et al. 2020; Bai et al. 2007; Zhao et al. 2012).

### Concordance between drug resistance phenotype and genotype-based predictions

Our analysis has also provided an assessment of the accuracy of genotype-based drug resistance prediction within a Vietnamese population. Contextualising the results we obtained is difficult, as prior studies have typically relied on much smaller sample sizes (Kohl et al. 2018; Schleusener et al. 2017; Macedo et al. 2018), have not reported data for the same subset of drugs analysed here (Zignol et al. 2018), or have systematically removed isolates with uncharacterised mutations in target genes (Cryptic Consortium, 2018). We contrast our results here with those of Mahe et al. (2019), who analyse a large sample (>6,000 isolates) using the same prediction catalogue and methodology.

Our sensitivity values for INH and STR prediction are comparable with those of Mahe et al. (2019) (0.89 and 0.77 respectively), and specificity values for all four first-line drugs are equal to or higher than those of Mahe et al. (2019) (INH=0.97; RIF=0.98; EMB=0.93; STR=0.91). Our sensitivity values for the two drugs with the lowest levels of phenotypic resistance, RIF and EMB are, however, lower than those of Mahe et al. (2019) (0.91 and 0.93 respectively). Also, our ability to predict resistance to second-line drugs was considerably lower than Mahe et al. (2019), who report sensitivity values of 0.92, 0.92 and 0.89 for KAN, AMI and aggregated Fluoroquinolones respectively.

We conclude that prediction accuracies for the two drugs with the highest rates of resistance, INH and STR, are comparable with global benchmarking standards. We also note that the frequency distributions of markers explaining resistance in correctly predicted isolates are consistent with a recent survey of the Vietnamese population (Mai et al. 2018), and the recently published WHO catalogue (WHO, 2021a). In all instances, the most commonly reported resistance conferring variants in our cohort were also amongst the most frequent in the catalogue collated by the WHO (2021a).

A future avenue of research may involve investigating the genetic basis of resistance in the high number of EMB resistant isolates which were predicted to be susceptible using the TBProfiler catalogue (N=42/75, 56%). It is clear that large studies correlating genotypic predictions of resistance with clinical outcomes are essential to improve the accuracy of genotypic resistance prediction and to resolve discrepancies between phenotypic and genotypic prediction, especially for the antimycobacterial drugs with poorly characterised resistance mechanisms.

### High rates of acquisition and transmission of drug resistance in Beijing lineage isolates

This investigation has explored the dynamics of *Mtb* drug resistance evolution in a high burden South-East Asian setting. Analysis of cohorts from other geographical regions have alternately emphasized the roles of transmission (Woolenberg et al. 2017; Casali et al. 2014; Yang et al. 2017) or *de novo* acquisition (Cohen et al. 2015; Ektefaie et al. 2021; Manson et al. 2017) in explaining rates of drug resistance. The drug resistance burden in our Vietnamese cohort was explained by both the *de novo* acquisition of resistance, and ongoing transmission of resistant isolates. As can reasonably be expected, the rate of transmitted INH resistance reported here (63.6%) is lower than reported in Woolenberg et al. (2017) (90%), who explore an outbreak within a small Russian population, yet higher than values reported by Ektefaie et al. 2021 (∼40%) who analyse a diverse global cohort.

Importantly, we find that drug resistance burden in Vietnam is predominantly driven by Beijing lineage isolates, which develop resistance conferring mutations at a faster rate than other isolates, and display a tendency to transmit resistance between hosts. These findings contribute to our understanding of the biology and life history of Beijing lineage *Mtb* isolates, which have previously been shown to display enhanced virulence and transmissibility than isolates from other lineages (Holt et al. 2018). This data therefore highlights an additional danger associated with the Beijing lineage, which is gradually supplanting endemic lineage 1 isolates within Vietnam (Holt et al. 2018).

### Limitations of this analysis

A caveat which must be applied to our findings is that our phylogenetic methods are unable to model the evolution of resistance in isolates for which their phenotype cannot be explained by markers in the TB-Profiler catalogue. It is possible that inferences surrounding the evolution of RIF resistance, for instance, may change with the identification of more resistance conferring markers. Resistance explained by indels is similarly unable to be modelled using our methods, however, this class of variant represents a small fraction of our dataset.

### Implications for TB control in Vietnam

In addition to highlighting dynamics of drug resistance evolution within our cohort, we have also established that resistance to INH and STR arose earlier, on average, than resistance to RIF, and is now more widespread within the Vietnamese population. The inference of early INH resistance evolution is consistent with prior studies, and mimics results obtained from a South African population (Cohen et al. 2015), and two surveys of global *Mtb* isolates (Manson et al. 2017; Ektefaie et al. 2017).

We conclude, as do Manson et al. (2017), that a rapid assay for INH resistance will allow the detection of ‘pre-MDR’ TB, and offer the high number of patients with INH resistant TB, treatment options which include other drugs. Unlike the cohort analysed by Manson et al. (2017), however, we find that a high percentage of RIF resistance isolates are susceptible to INH (20.7%). This finding further supports the utility of an INH resistance assay, which may be applied after a patient tests positive for RIF resistance using the MTB/RIF Xpert test. Several WHO approved tests for INH resistance could be implemented (WHO, 2016). Application of such INH resistance tests will allow the 20% of RIF resistant cases which remain susceptible to INH to be treated with an INH-containing regimen.

Finally, the high rates of resistance to a number of second line drugs highlights the utility of WGS-based individualised therapy for drug resistant TB in Vietnam. The poor sensitivity values calculated, however, stresses the need to develop a greater understanding of the genetic variants implicated in resistance to these drugs.

## Materials and Methods

### Study population

Bacterial isolates (N=2,619) from patients with pulmonary TB (PTB) (N=2,446) and tuberculous meningitis (TBM) (N=173) were collected as part of larger clinical studies (Heemskerk et al. 2016; Thai et al. 2018; Thwaites et al. 2004). Patients with PTB were defined as HIV-negative adults (>18 years) sputum culture positive for *M. tuberculosis*. Isolates (N=1,654) were collected in 8 district TB units (DTUs) in Ho Chi Minh City (HCMC), Vietnam between December 2008 and July 2011 (Thai et al. 2018). A further 792 isolates from PTB patients were similarly collected at these DTUs in HCMC as an extension to this clinical study, from 2011-2013. Patients with TBM were defined as HIV-negative patients, >15 years old, with cerebral spinal fluid (CSF) culture positive *Mtb* and were recruited into two clinical trials conducted at the Hospital for Tropical Diseases and Pham Ngoc Thach Hospital for TB and Lung Diseases in HCMC (Heemskerk et al. 2016; Thwaites et al. 2004). 62 were collected between 2001-2003 (Thwaites et al. 2004) and 111 were collected between 2011-2015 (Heemskerk et al. 2016), with a total of 173 isolates from TBM patients included in this genomic study.

### Ethical approvals

All study protocols for PTB and TBM studies were approved by the Institutional Research Board of Pham Ngoc Thach Hospital for TB and Lung Disease, HCMC Health Services and the Oxford Tropical Research Ethics Committee, UK. Approval for the genomics study was granted from the Health Sciences Human Ethics Sub-Committee at the University of Melbourne, Australia (ID:1340458). Written informed consent was obtained from all patients.

### Phenotypic drug susceptibility testing (DST)

Phenotypic DST was performed using two techniques. In the first method, isolates were subcultured in Mycobacterial Growth Indicator Tubes (MGIT) for phenotypic DST on the first line drugs, INH 0.1μg/ml; STR 1.0μg/ml; RIF 1.0μg/ml; EMB; 5.0μg/ml, using the BACTEC MGIT 960 SIRE Kit (Becton Dickinison) according to the manufacturer’s instructions. The second method used the UKMYC5 plate designed by the CRyPTIC consortium which enables minimum inhibitory concentration (MIC) measurement for 14 different anti-tuberculosis compounds. The UKMTC5 MIC plate method was used as described in Rancoita et al. (2018). The critical concentrations used to determine drug resistance were BDQ 1.0μg/ml; KAN 2.5μg/ml; ETH 5.0μg/ml; AMI 1.0μg/ml; EMB 5.0μg/ml; INH 0.1μg/ml; LEV 1.0μg/ml; MXF 0.5μg/ml; DLM 0.12μg/ml; LZD 1.0μg/ml; CFZ 1.0μg/ml; RIF 1.0μg/ml; RFB 1.0μg/ml; PAS 4.0μg/ml.

### DNA extraction and sequencing

Lowenstein Jensen media was used to subculture isolates at the Oxford University Clinical Research Unit, Vietnam, prior to DNA extraction using the cetyl trimethylammonium bromide extraction protocol as described previously (Caws et al. 2008). DNA was shipped to the University of Melbourne (UoM) (N=1,827) and the National University of Singapore (NUS) (N=792) for whole genome sequencing. At GIS, genomic DNA was first quantified by Picogreen assay, followed by shearing using the Covaris. Library preparation was done using a commercially available kit, NEBNext® Ultra™ DNA Library Prep Kit for Illumina® following the manufacturer’s protocol. The quality of the libraries were QC via LabChip GX or Agilent D1000 ScreenTape before pooling. After pooling, the pooled library was QC using Agilent high sensitivity DNA kit and KAPA quantification before sequencing on the Illumina HiSeq 4000 (Illumina, San Diego).

### *Mtb* genome data and SNP calling

*Mtb* genome data from a subset of PTB patients have been previously described and is denoted here as the ‘published subset’ (Holt et al. 2018). These genomes are available at the European Nucleotide Archive (accession ID PRJNA355614; https://www.ebi.ac.uk/ena/browser/view/PRJNA355614). The remaining *Mtb* genome data will be available via the NCBI database upon publication of this manuscript. The complete genome collection is denoted as the HCMC PTB/TBM genome dataset. Variant calling for *Mtb* isolates was carried out using the RedDog pipeline V1beta.11 (https://github.com/katholt/RedDog) with default settings, which uses BowTie (Langmead et al. 2009) for read mapping and Samtools (Li, 2011) for variant calling. After variant calling, samples for which less than 90% of their reads mapped to the *Mtb* H37Rv reference genome (NC_000962.3), and with high proportions of heterozygous sites were removed. Variants called in repetitive regions, as defined by Holt et al. (2018), were also removed. *Mtb* lineages and sublineages were assigned using fast-lineage-caller (Freschi et al. 2021) on per-sample vcf files, using the scheme of Coll et al. (2014). Sequence reads from an isolate belonging to lineage 5 were also incorporated into the above variant calling pipeline as an outgroup for all subsequent phylogenetic analysis.

### Drug resistance prediction using whole genome sequencing data

Whole genome sequencing-based resistant prediction was performed with TB-Profiler v4.2.0 (Phelan et al. 2019) using the most up to date database available (16/02/2022). Read data were screened for mutations associated with resistance to 4 first-line drugs, (INH, RIF, EMB and STR), and second-line drugs for which phenotypes were available, and which are covered by the TB-Profiler catalogue.

### Phylogenomic analysis

We inferred a phylogeny from 2,542 *Mtb* genomes using RAxML v8.2 (Stamatakis, 2014), using a GTR model of nucleotide substitution. Ancestral state reconstruction was then performed using SNPPar v1.0 (Edwards et al. 2021) with default settings. Biallelic SNPs conferring drug resistance were defined according to TB-Profiler, and extracted from the SNPPar output. All downstream analyses of tree diversity metrics were carried out using custom Unix and R scripts (R Core Team, 2021), and the R packages ape v5.6.2 (Paradis & Schliep, 2018), treeio v1.18.1 (Wang et al. 2019) and phangorn v2.8.1 (Schliep, 2010).

To make inferences regarding the dynamics of drug resistance evolution and transmission, we followed the theory applied by recent studies (Manson et al. 2017; Ektefaie et al. 2021; Woolenberg et al. 2017). Mtb isolates for which resistance mutations were inferred to have arisen on a terminal branch of the phylogeny were taken to indicate putative cases of acquired resistance. These mutations were plotted as blue points in Figure 3a on the tip of the phylogeny downstream of the branch on which they arose.

We interpreted resistance mutations which could be localised to nodes of the phylogeny, and which are therefore inherited by multiple descendant tips, to indicate cases of transmitted resistance. These mutations were plotted as red points in Figure 3a on the node downstream of the branch on which they arose. The small number of isolates (N=5) across the phylogeny which developed resistance-conferring variants on their terminal branch, but which had already descended from a resistant node, were classified as cases of transmitted resistance.

We excluded EMB from this analysis as our ability to accurately model transmission dynamics would be limited due to the low predictive power of EMB resistance variants (59.8% of isolates with putative EMB resistance variants were phenotypically susceptible; Figure 2a), in addition to the low sensitivity demonstrated previously (Figure 2a).

### Statistical analysis

Analyses were done using R version 4.1.1 and two-sided P<0.05 was considered statistically significant. The overall lineage distribution of *Mtb* isolates across gender, clinical phenotype and resistance to the first and second anti-TB drugs was presented as number and percentage (%). Pearson’s χ^2^ test was performed to investigate the relationship between categorical variables, including lineages and other clinical characteristics, naming gender, clinical phenotype and drug resistance. It was also utilized to assess the relationship between rates of drug resistance and lineages, as well as resistance and transmission ability. Mann-Whitney U test was employed to test the difference for non-normally distributed variables among lineages, including count of total mutations relative to the number of resistance conferring mutations and lengths of terminal branches leading to resistance. The association between age and lineage (lineage 1 and 2) was assessed using linear regression. To investigate the prediction accuracy for drug resistance using WGS data we calculated sensitivity, defined as the proportion of resistant isolates correctly predicted; specificity, defined as the proportion of susceptible isolates correctly predicted; positive predictive value (PPV) and negative predictive value (NPV).

## Data Availability

Mtb genome data from a subset of PTB patients have been previously described. These genomes are available at the European Nucleotide Archive (accession ID PRJNA355614; https://www.ebi.ac.uk/ena/browser/view/PRJNA355614). The remaining Mtb genome data will be available via the NCBI database upon publication of this manuscript.

## Acknowledgements

We would like to thank the clinical staff who recruited patients into our study from the following District TB Units (DTUs) in HCMC, Vietnam: Districts 1, 4, 5, 6 and 8, Tan Binh, Binh Thanh and Phu Nhuan; and also our colleagues from Pham Ngoc Thach Hospital for Tuberculosis and Lung Disease, HCMC Vietnam. We would like to acknowledge the Cryptic consortium, http://www.crypticproject.org/, for providing the UKMYC5 plates for DST. This work was supported by the National Health and Medical Research Council, Australia (Investigator grant APP1172853 to SJD); NHMRC (APP1056689) /A*STAR (12/1/21/24/6689) joint call to SJD/YYT; the Wellcome Trust UK (research training fellowship 081814/Z/06/Z to MC), (intermediate fellowship 206724/Z/17/Z to NTTT) and as part of their Major Overseas Program in Vietnam (089276/Z/09/Z to JF and 106680/B/14/Z to GT). The National University of Singapore Yong Loo Lin School of Medicine Aspiration Fund (NUHSRO/2014/069/AF-New Idea/04) contributed funding to the whole genome sequencing conducted in this study. MI was supported by the Munz Chair of Cardiovascular Prediction and Prevention at the Baker Heart and Diabetes Institute and the NIHR Cambridge Biomedical Research Centre (BRC-1215-20014) [*]. *The views expressed are those of the author(s) and not necessarily those of the NIHR or the Department of Health and Social Care.

