## Supplementary figures and images for "Rapid Acquisition and Transmission of Drug Resistance Amongst Beijing Lineage *Mycobacterium tuberculosis* in Vietnam"

### Figure S1

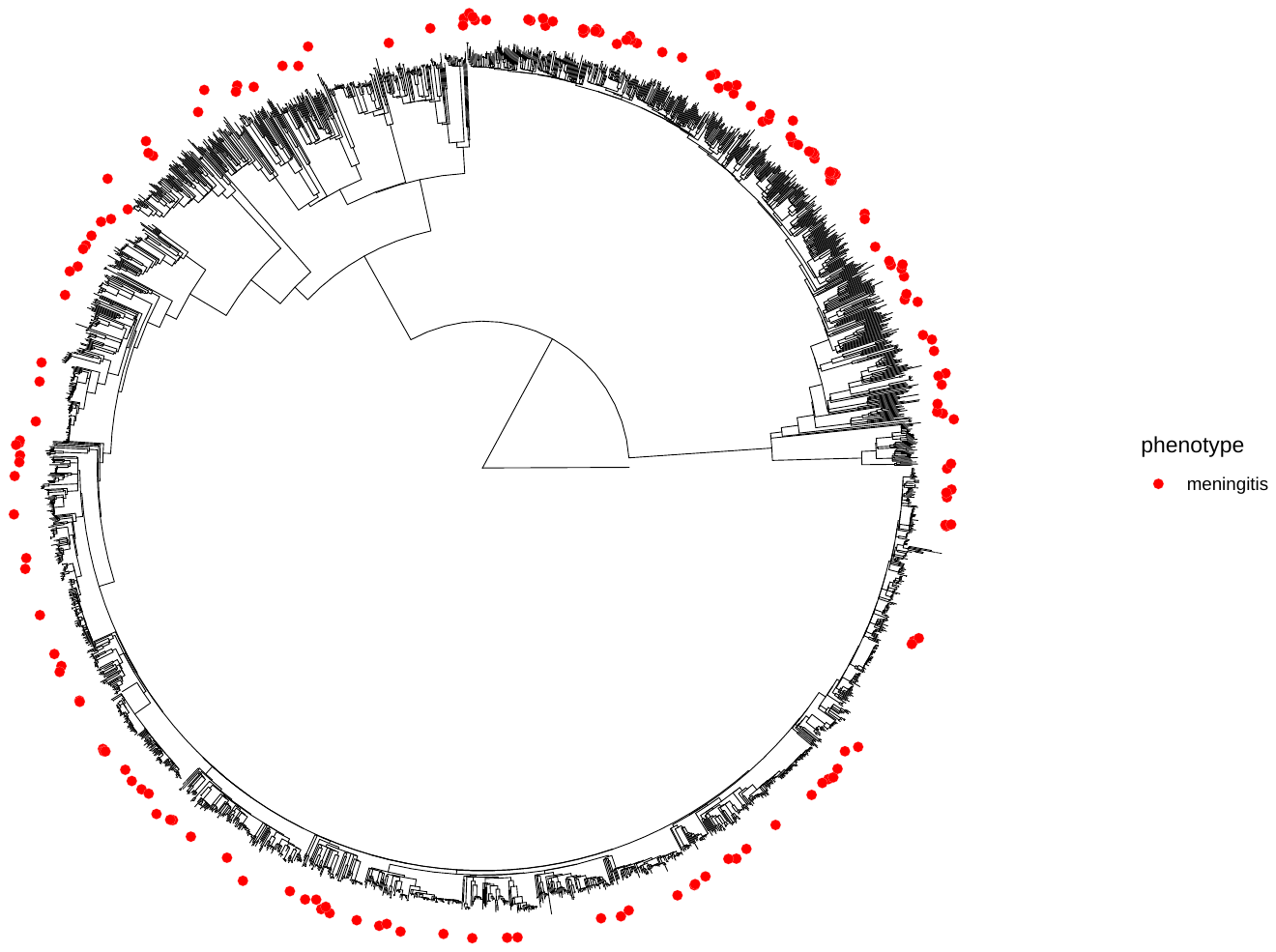

**Figure S1. Distribution of TBM across the phylogeny of N=2,542 Mtb isolates from HCMC, Vietnam.**
